# Time-varying consideration of health behaviours explains over 90% of the inequality in mortality associated with socioeconomic status

**DOI:** 10.1101/2024.10.01.24314725

**Authors:** Rebecca Harounoff, Danyal Sarwar, Maria Perez-Ortiz, Eric Brunner, John Shawe-Taylor

## Abstract

**Background:** Applying survival analysis techniques to epidemiological inference within research into ageing offers opportunities to estimate the association between exposure and outcome in longitudinal data. This study used Cox regression to investigate how socioeconomic inequality in mortality can be explained by exposure to various factors including smoking, diet, alcohol and physical activity. This study seeks to complement and extend previous work which found that the contribution of the socioeconomic gradient to inequalities in health was underestimated by baseline analysis.

**Methods:** Data was obtained from Whitehall II, a British longitudinal cohort study, which investigated social determinants of health. Analysis is based on 11 waves of data collected over 32 years on 10,308 civil servants aged between 35 and 90. Socioeconomic position was defined by baseline employment grade (1-3). During the follow-up 2,427 participants died. Extensive experimental analysis was conducted using a vast number of health behaviours. Cox regression produced an age-and-sex-adjusted hazard ratio for the socioeconomic inequality in mortality. Health behaviours (smoking, physical activity, alcohol consumption, and diet) were then added as covariates to determine the extent to which they statistically explain this inequality, and how this differed from the last similar analysis from 2009. This was done at baseline and longitudinally. The health behaviours were then combined linearly, nonlinearly and new health behaviours were added.

**Results:** Adding the above health behaviours as covariates statistically explained the socioeconomic gradient in mortality at baseline from 42% to 2009, to 51% to 2021. Longitudinal consideration increased the explanatory power, when all health behaviours were added as time-varying covariates, from 51% to 87%. Adding more variables in the form of a more comprehensive diet score statistically explained the gradient further, to 91%. The nonlinear model of smoking and exercise most accurately predicted mortality and had a 13% higher explanatory power when explaining the gradient compared to the linear model in longitudinal data.

**Conclusion:** In the Whitehall II study, socioeconomic position and mortality showed an association. There is a gain in explanatory power of the set of health behaviours at baseline when follow-up is extended by 12 years, from 42% to 51%. When changes in behaviour over the 32 years of follow-up were also accounted for, this association was now significantly explained by over 90%, compared with 51% when considered at baseline. We suggest that reverse causation is partly responsible for the almost complete explanation of the social gradient in mortality by health behaviours. These results would therefore lead us to question why health behaviours are socially patterned in the way that has been observed, which would be significant for targeting health behaviours in lower socioeconomic statuses.

## Introduction

### Background

With increasing life expectancy have come ageing populations [1]; it is expected that by 2050, one in six people will be over 65 [2]. This has heightened desire to understand the factors impacting longevity and quality of life in old age [3], which are complex and heterogeneous between individuals: encompassing lifestyle, psychosocial and disease-related factors [4].

Socioeconomic position can be said to impact on these factors, perhaps indicating why it shows a gradient in mortality: the gap in life expectancy in England between the 5^th^ and 95^th^ percentiles of earners is six years [5]. As well as having shorter lives, those living in poorer areas spend longer in poor health [5]. Investing in prevention has been shown to reduce overall health costs and welfare benefits [6].

Previous studies have aimed to investigate the contributions of health behaviours to social inequalities in mortality [7]. Health behaviours are crucial determinants of morbidity and mortality including cardiovascular disease and cardiometabolic risk worldwide [8][9]. A recent study established that ‘one in three premature deaths are attributable to socioeconomic inequalities’ [10].

However, important gaps remain. Typically, health behaviours are assessed at one point in time (baseline), which only partially reflects the inequality between socioeconomic status and mortality. A person’s health behaviours are unlikely to remain constant throughout the life course. Additionally, these papers do not consider nonlinear relationships that may exist between variables. These will be studied further in this work.

Previous work such as in *Stringhini et al*. [7] investigated the influence of health behaviours (smoking, alcohol consumption, diet, physical activity) on the association between socioeconomic position and mortality. These statistically explained 42% of the gradient in mortality at baseline. When considered longitudinally at several phases over 24 years, however, these behaviours explained 72% of this inequality [7].

### Objectives of this work

**Aim 1:** To replicate results obtained by *Stringhini et al*. [7] with follow-up to 2009 to validate the proposed Cox model. To then expand this to incorporate follow-up to present day (32 years) for the baseline model.
**Aim 2:** To build a longitudinal model to account for behavioural changes over follow-up
**Aim 3:** To expand the model to include a richer combination of variables.
**Aim 4:** To examine the nonlinear relationship between smoking and exercise. Previous studies have investigated the relationship between smoking and exercise and have found that smoking was associated with decreased exercise [11]. Even among the young, smoking is detrimental to physical activity. However, these studies have not investigated how a nonlinear relationship between these variables may affect the inequality in mortality associated with socioeconomic status.

**Main outcome measure:** All-cause mortality

### Ethical approval

Ethical approval was obtained under the work conducted by Eric Brunner’s group at UCL School of Life and Medical sciences.

REC reference: 85/0938 IRAS project ID: 142374.

Data was accessed through the Dementias Platform UK (a medical research council) secure platform. The dataset contains a vast amount of confidential information, therefore, data cannot be imported or exported from the platform.

## Methods

This section explains the characteristics of the study population and how it was prepared, including the approach to missing data for analysis, as well as evaluating the statistical methods used.

### Study population

Data was obtained from Whitehall II a British longitudinal cohort study, which investigated social determinants of health [12]. Civil servants were invited to participate in this study via letter, 73% consented in writing giving a total of 10,308 civil servants aged between 35 and 90. Whitehall II data from phases 1 to 11 was used (spanning 1985-2017), with mortality data to 2021. Only data from odd phases was used: these were the phases when clinical examination was undertaken in addition to questionnaire completion.

Socioeconomic position was estimated from employment grade at baseline. This variable takes three values: 1, lower grade (clerical/support roles); 2, intermediate grade (professional/executive roles); 3, higher grade (administrative roles).

### Health behaviours

Alcohol consumption in units was calculated as a combination of units of beer, wine, and spirits consumed in a week, and scoring allocated analogously to the scoring of *Stringhini et al*. [7]: (1) never (0 unit/week); (2) moderate (1-21 units/week for men, 1-14 for women); (3) heavy (>21 units/week for men, >14 for women) [13].

For the purpose of comparison, diet was scored in accordance with the scoring devised by *Stringhini et al*. [7], where participants were classified as (1) unhealthy if participants ate white bread most frequently, consumed whole milk, and ate fruit and vegetables less than 3 times per month; (2) healthy if they ate wholemeal, wheatmeal, or other brown bread most frequently, did not consume milk or only used skimmed or other types of milk, and ate fruit and vegetables daily or 2 or more times per day; (3) moderately healthy if their dietary pattern was in between these 2 descriptions [9].

Physical activity was assessed based on hours per week of moderate and vigorous physical activity, and scored as by *Stringhini et al*. [7]: (1) active (>2.5 hours/week of moderate physical activity or >1 hour/week of vigorous physical activity); (2) inactive (<1 hour/week of moderate physical activity and <1 hour/week of vigorous physical activity); (3) moderately active if their physical activity fell between these 2 descriptions [13].

### Removal of missing data

Rows corresponding to participants with missing data values were removed in Python using the *Pandas* module. 93% of participants did not have missing data values, motivating this decision. For comparison, *Stringhini et al*. [7] removed all rows corresponding to missing data in an analogous way.

### Survival analysis and Cox regression

Where time-to-event is the main outcome under analysis (as in this study), it is referred to as *survival time*, which may differ between participants. Indeed, some- the majority in this study-may not undergo the event at all and thus have no true time-to-event. This is known as data censoring. This necessitates unique methods- the methods of *survival analysis* [14].

Cox regression is one of the most used methods in survival analysis because a hazard ratio (HR) can be calculated, and the estimated hazards are always non-negative [15]. The coefficients in the Cox regression relate to a hazard, where a hazard ratio above 1 implies a covariate is positively associated with the event occurring. This motivates Cox regression for our analysis, where our coefficients would indicate the extent to which health behaviours influenced the gradient in mortality.

#### Baseline Cox

The univariate Cox model uses data at a single point in time. In this case, we use health behaviour data from baseline. This method is used so we can compare how the socioeconomic gradient in mortality may change when data is assessed longitudinally. The central analysis was achieved using Stata statistical software, version 17 to perform Cox regression standardising for age and sex. The outputted hazard ratios corresponded to the relative risk of mortality of that socioeconomic group (1, 2, or 3) compared to the highest group, adjusted for age and sex. This would then be the hazard ratio unadjusted for health behaviours. Health behaviours were then added as covariates *x*i in turn to generate adjusted hazard ratios.

#### Longitudinal Cox

The longitudinal Cox model incorporates data from different phases, with further years of follow up data in comparison to Stringhini. It allows us to quantify the effect of repeated measures of covariates on the effect of mortality. Where individuals had follow-up data available, the covariates for health behaviours may change with time and thus were time-dependent covariates [16], whereas socioeconomic status remained as a time-fixed exposure at baseline. Where participants had missing data on health behaviours at one of the follow-up assessments this was substituted, where available, with data from the phase directly prior or subsequent. Phases after a participant’s death were removed. The Cox regression was then run analogously to the above, now providing insight into how changes in health behaviours across follow-up influenced the socioeconomic gradient in mortality.

#### Percentage attenuation

The percentage attenuation was calculated using the formula below [7] and was used for both baseline and longitudinal cox regression models. This allows us to quantify the role of each health behaviour in explaining the socioeconomic inequality statistically in a conservative way using a log scale. *ln*(HR_un*adjusted for* h*ealt*h *be*h*aviours*_) − *ln*(HR_*adjusted for* h*ealt*h *be*h*aviours*_) × 100 *ln*(HR _un*adjusted for* h*ealt*h *be*h*aviours*_)

## Results

**Aim 1: To replicate results obtained by *Stringhini et al*. [7] with follow-up to 2009 to validate the proposed Cox model. To then expand this to incorporate follow-up to present day (32 years) for the baseline model**.

The same methods as used by *Stringhini et al*. [7] for mortality data to 2009 were applied to data up to 2021 to investigate any changes in hazard ratios when applied over a longer time period. This meant more deaths: 654 deaths to 2009, increasing to 2427 to 2021, from 9590 participants without missing data values. The baseline hazard ratios are compared in Table 1.

**Table 1:** Role of health behaviours in statistically accounting for the association between socioeconomic position and all-cause mortality^b^: comparison at baseline between data from 2009 and from 2021.

|  | Baseline Hazard Ratio 2009<br>(Stringhini et al) |  | Baseline Hazard Ratio 2021 |  |
| --- | --- | --- | --- | --- |
|  | HR (95% CI) | % Attenuation<sup>c</sup> | HR (95% CI) | % Attenuation<sup>c</sup> |
| **Model 1<sup>a</sup>** | 1.60<br>(1.26 to 2.04)<sup>d</sup> | N/A | 1.55<br>(1.37 to 1.76) | N/A |
| **Plus smoking** | 1.36<br>(1.06 to 1.74) | 32 | 1.33<br>(1.17 to 1.51) | 35 |
| **Plus alcohol consumption** | 1.58<br>(1.24 to 2.03) | 3 | 1.51<br>(1.34 to 1.72) | 6 |
| **Plus diet** | 1.55<br>(1.21 to 1.98) | 7 | 1.47<br>(1.26 to 1.69) | 12 |
| **Plus physical activity** | 1.57<br>(1.23 to 2.00) | 5 | 1.49<br>(1.31 to 1.69) | 9 |
| **Fully adjusted** | 1.31<br>(1.02 to 1.69)<sup>e</sup> | 42 | 1.24<br>(1.08 to 1.41) | 51 |

Abbreviations: CI, confidence interval; HR, Hazard ratio; NA data not applicable
a: HR adjusted for age and sex only
b: In the Whitehall II study, there were 654 deaths out of a total of 9590 in 2009 and 2427 deaths in 2021
c: Percentage attenuation= $100 \times (\ln_{\text{Model 1+ health behaviours}} - \ln_{\text{Model 1}}) / (\ln_{\text{Model 1}})$
d: Lowest socioeconomic position compared to highest, adjusted for sex and age
e: Includes all listed health behaviours

The hazard ratio for the unadjusted model represents the difference in hazard rate between individuals of the lowest and highest employment grade when adjusted for age and sex only. The listed health behaviours were then added individually as covariates and adjusted hazard ratios between lowest and highest employment grades calculated, and then all behaviours added simultaneously. The associated percentage attenuations were then calculated (which indicate the extent to which a health behaviour explains the socioeconomic gradient in mortality). Results on the mediating role of health behaviours compared at baseline with *Stringhini et al*. [7] (2009) and our analysis (2021) is visible in Table 1.

Comparing follow-up to 2009 and to 2021, in the first model adjusting only for age and sex at baseline, the HR decreased slightly from 1.60 (95% CI 1.26-2.04) to 1.55 (95% CI 1.37-1.76) from 2009 to 2021. The explanatory power of the health behaviours at baseline increases when follow-up time increases. This is clear from the increased percentage attenuation for all listed behaviours.

In addition, the confidence intervals have narrowed significantly, possibly due to several more individuals reaching the time-to-event (mortality) between 2009 and 2021. The increase in deaths provides a higher statistical power which generates increased confidence in the explanatory power of the model. Overall, health behaviours at baseline can now be said to account for 51% of the gradient in mortality.

**Aim 2: To build a longitudinal model to account for behavioural changes over follow-up**

Following on from this, it was investigated how hazard ratios considering changes in health behaviours across the period of follow-up compared with those solely assessing health behaviours at baseline (above). These longitudinal analyses, where the health behaviours were entered as time-varying covariates, are shown in Table 2, with the percentage attenuation again alongside the hazard ratios. These percentage attenuations were then plotted (Figure 2) against those obtained above from assessment solely at baseline, to show how the statistical explanation differs.

**Table 2:**
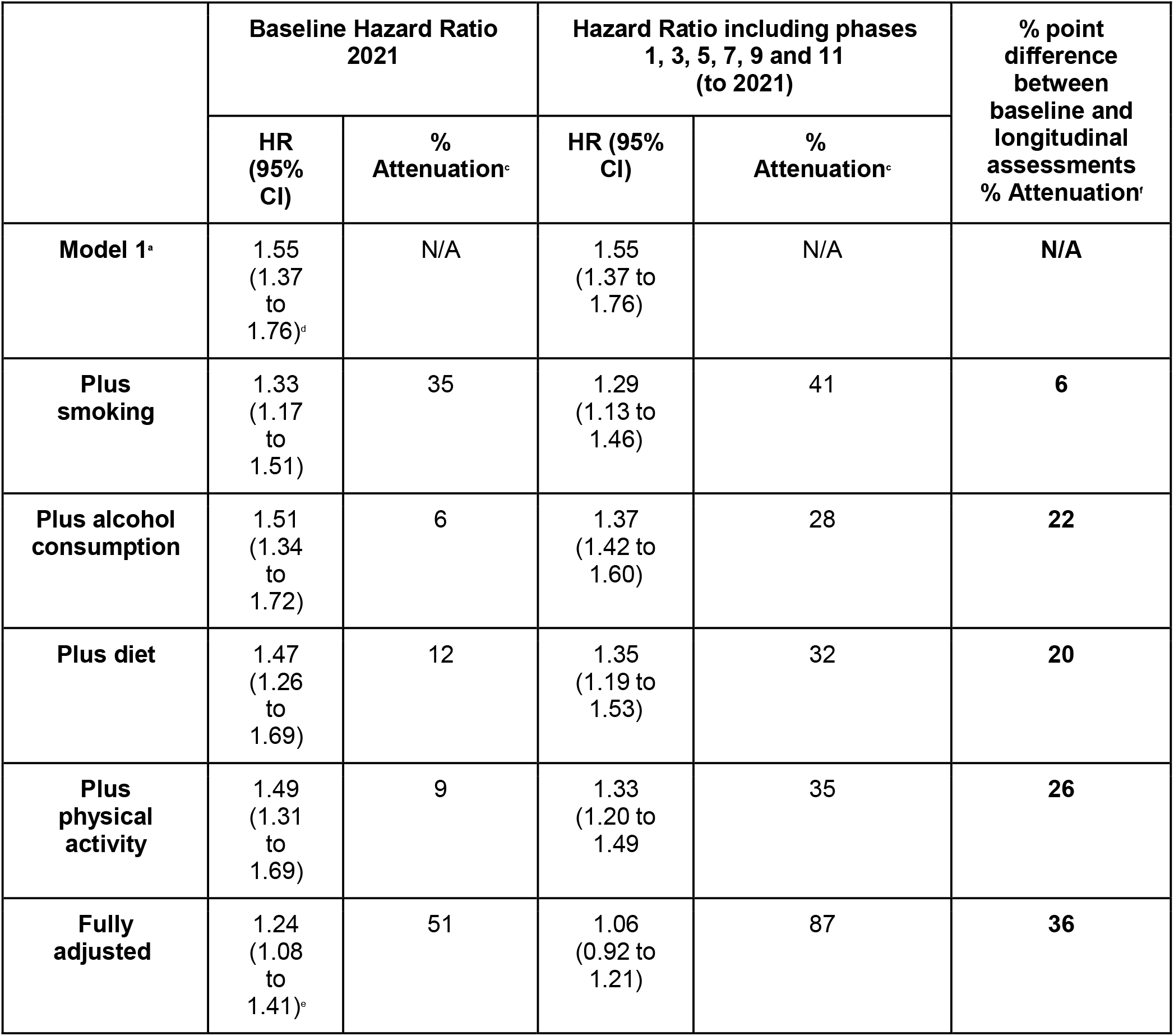
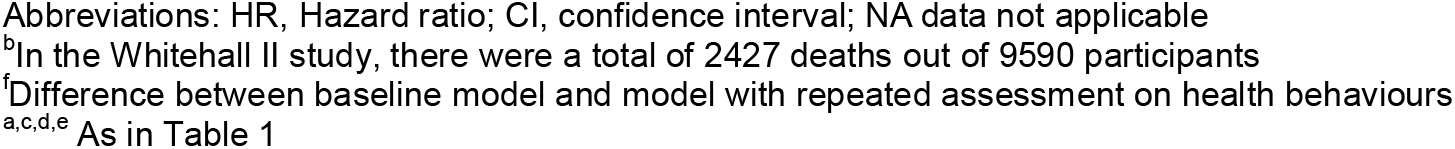
Role of health behaviours in statistically accounting for the association between socioeconomic position and all-cause mortality^b^: comparison of baseline and longitudinal up to and including 2021.

|  | Baseline Hazard Ratio 2021 |  | Hazard Ratio including phases 1, 3, 5, 7, 9 and 11 (to 2021) |  | % point difference between baseline and longitudinal assessments % Attenuation<sup>f</sup> |
| --- | --- | --- | --- | --- | --- |
|  | HR (95% CI) | % Attenuation<sup>c</sup> | HR (95% CI) | % Attenuation<sup>c</sup> |  |
| **Model 1<sup>a</sup>** | 1.55<br>(1.37 to 1.76)<sup>d</sup> | N/A | 1.55<br>(1.37 to 1.76) | N/A | **N/A** |
| **Plus smoking** | 1.33<br>(1.17 to 1.51) | 35 | 1.29<br>(1.13 to 1.46) | 41 | **6** |
| **Plus alcohol consumption** | 1.51<br>(1.34 to 1.72) | 6 | 1.37<br>(1.42 to 1.60) | 28 | **22** |
| **Plus diet** | 1.47<br>(1.26 to 1.69) | 12 | 1.35<br>(1.19 to 1.53) | 32 | **20** |
| **Plus physical activity** | 1.49<br>(1.31 to 1.69) | 9 | 1.33<br>(1.20 to 1.49) | 35 | **26** |
| **Fully adjusted** | 1.24<br>(1.08 to 1.41)<sup>e</sup> | 51 | 1.06<br>(0.92 to 1.21) | 87 | **36** |

Abbreviations: HR, Hazard ratio; CI, confidence interval; NA data not applicable
<sup>b</sup>In the Whitehall II study, there were a total of 2427 deaths out of 9590 participants
<sup>f</sup>Difference between baseline model and model with repeated assessment on health behaviours <sup>a,c,d,e</sup> As in Table 1

**Figure 1:**
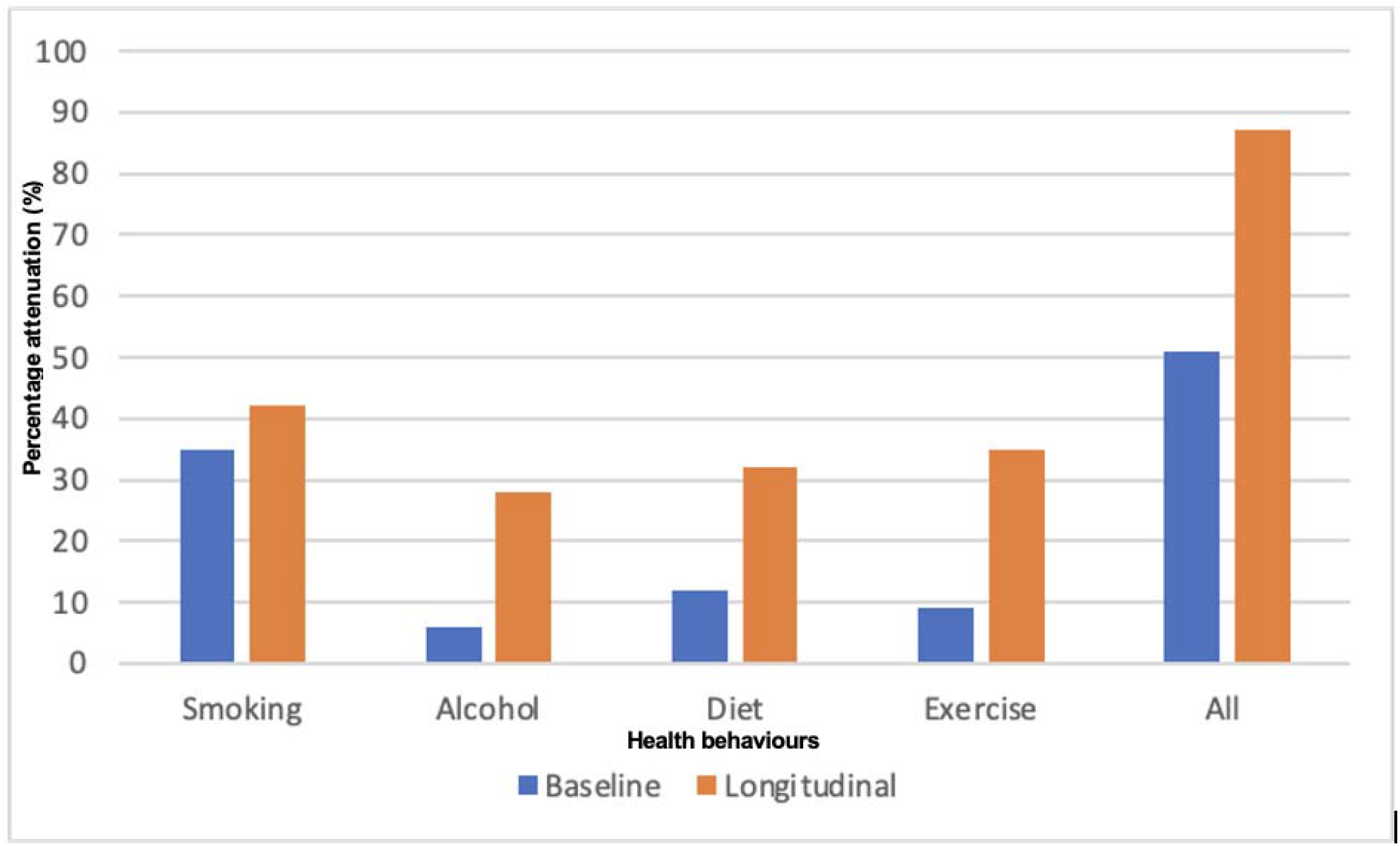
Comparison of health behaviours assessed at baseline and longitudinally.

**Figure 2:**
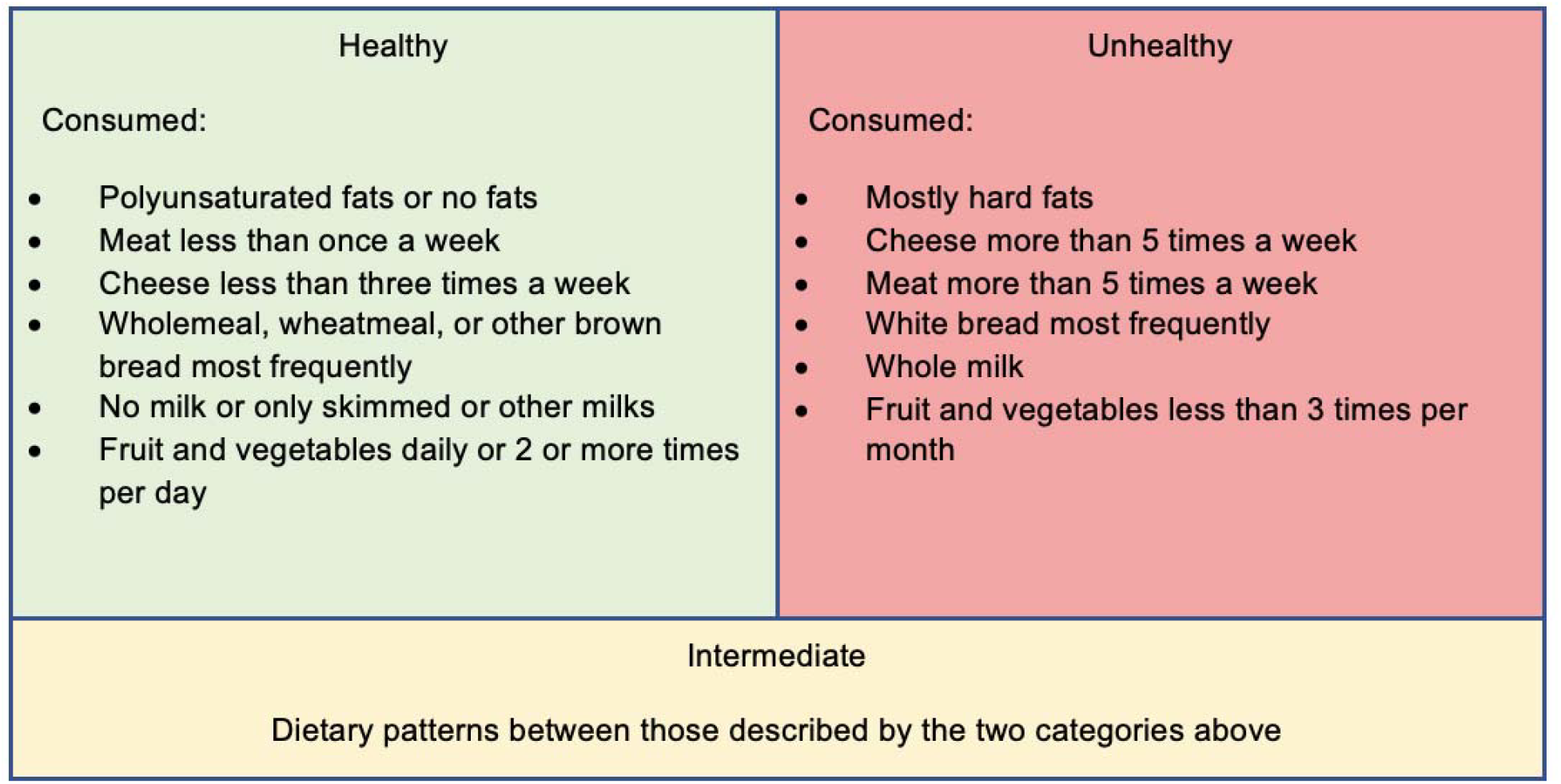
New diet scoring system explained considering other factors included in the NHS Eatwell Guide [17].

The attenuation increase for the time-varying exposure model compared to the baseline model was modest (35 to 41%) for smoking compared to diet (12 to 32%), physical activity (9 to 35%), and alcohol consumption (6 to 28%). This implies that the explanatory power of smoking towards the gradient in mortality associated with employment grade was not substantially increased when entered as a time-dependent covariate, unlike for diet, physical activity, and alcohol consumption. More broadly, a large majority (87%) of socioeconomic inequality in mortality is now explained when all these health behaviours are examined together and treated as time-dependent covariates. Overall when considering the above health behaviours smoking has the largest contribution, followed by exercise, diet and alcohol.

**Aim 3: To investigate the effect of incorporating more variables, in this case by forming a more detailed diet score, on the results obtained by the model**.

In the analyses above, the number of variables used in the scores for behaviours was low compared to the number available. Diet was classified using a scoring metric devised by *Stringhini et al*. [7] which incorporated only the type of bread and milk, and amount of fruit and vegetables consumed. To consider the combined effect of more variables on the hazard ratio, a new diet score was formulated (Figure 2) where consumption of meat, cheese, and fat were added. This doubled the number of dietary factors considered, and now accounted for all food groups stipulated in the NHS Eatwell Guide [17]. The effect of this new diet score on the hazard ratios for the longitudinal analyses, and associated percentage attenuations, are displayed in Table 3.

**Table 3:** Effect on baseline hazard ratios for deaths to 2021 of incorporating more variables into the diet component of the health behaviours on the longitudinal analysis of their role in explaining the association between socioeconomic position and all-cause mortality^a^.

|  | Hazard Ratio including phases 1, 3, 5, 7, 9 and 11 (to 2021) |  |
| --- | --- | --- |
|  | HR (95% CI) | % Attenuation |
| Model 1 | 1.55<br>(1.37 to 1.76) | - |
| Plus Stringhini et al. diet scores<sup>a</sup> | 1.35<br>(1.19 to 1.53) | 32 |
| Plus new diet scores<sup>c</sup> | 1.30<br>(1.15 to 1.48) | 40 |
| Full adjustment<sup>b</sup> including Stringhini diet scores | 1.06<br>(0.93 to 1.22) | 87 |
| Full adjustment<sup>b</sup> including new diet scores | 1.04<br>(0.91 to 1.18) | 91 |

Abbreviations: CI, confidence interval; HR, Hazard ratio; NA data not applicable
<sup>a</sup>Of a total of 9590 participants, there were 2427 deaths until 2021
<sup>b</sup>All health behaviours considered, as in Table 1
<sup>c</sup>As in Table 1
<sup>d</sup>As in Figure 2

When entered as a time-varying covariate, the new diet scoring explains the socioeconomic gradient in mortality better than the previous scoring: by 40% vs 32%, a relative increase of 25%. Furthermore, when the new diet scores replace the old in the full adjustment along with the other health behaviours, 91% of this inequality in mortality is now accounted for: an overwhelming majority.

**Aim 4: To develop an extended Cox regression model by taking into consideration non-linear effects that exist in prognostic factors, specifically to test whether the non-linear relationship between smoking and physical activity may affect the inequality in mortality associated with socioeconomic status**

Table 2 shows smoking has the largest percentage attenuation; therefore, it is the largest contributor statistically among the modelled health behaviours to the socioeconomic inequality in mortality. Additionally, physical activity in the longitudinal data has the second-largest percentage attenuation. With this knowledge, the effect of smoking was combined with exercise to create a non-linear model to see, once this relationship is taken into account, how the inequality in mortality associated with socioeconomic status may change. In linear models each covariant is presented independently whereas in a nonlinear model we are suggesting that smoking and exercise are hierarchically connected. Some research has shown that smoking is detrimental to physical activity [18], whilst other researchers have shown that regular exercise may be effective in preventing the negative effects associated with smoking [19].

### Categorisation

Based on NHS guidelines to do at least 150 minutes of moderate-intensity exercise per week or 75 minutes of vigorous-intensity exercise per week, participants were classified as active and inactive. A non-linear model was created (using Python nested if statements) between smoking and exercise where participants were classified on a scale of 0-3. Smokers who don’t exercise were classified as 3, smokers who exercise 2, non-smokers who don’t exercise 1 and non-smokers who exercise 0. Participants with missing data were removed in Python. In the linear model smoking and exercise were both included simultaneously as independent covariates in the cox regression.

### Results

Of a total of 10,308 participants, 9,775 participants were included in the study and 533 were excluded based on missing data. A total of 2,480 deaths were recorded in the cohort between phases 1 and 11. This set of analyses included 186 more participants than the set of analyses conducted for Aim 1. This is because these participants had missing health behaviour data for diet or alcohol consumption, but not from hours of physical activity or smoking status. Table 3 compares the baseline Cox regression with the multivariate Cox regression as conducted previously. The C-index is included to measure the performance of each model respectively.

As is evident from Table 4 when a non-linear relationship between smoking and exercise is created in longitudinal data, the HR decreased significantly from 1.36 in the linear model between smoking and exercise to 1.28 in the nonlinear model. Therefore, it explains 13% more of the gradient that exists between lower and higher socioeconomic positions when using the nonlinear relationship instead of the linear relationship in the longitudinal cox regression. The model which included the nonlinear relationship between smoking and exercise using longitudinal data, has the highest C-index; this means that is the model which best predicts mortality on a test set of data.

**Table 4:** Role of the non-linear relationship between smoking and exercise in statistically accounting for the association between socioeconomic position and all-cause mortality^a^.

|  | Baseline Hazard Ratio<br>2021 |  |  | Hazard Ratio including<br>phases 1, 3, 5, 7, 9 and 11<br>(to 2021) |  |  |
| --- | --- | --- | --- | --- | --- | --- |
|  | HR (95% CI) | %<br>Attenuati<br>on<sup>b</sup> | C-Index<sup>d</sup> | HR (95% CI) | %<br>Attenuati<br>on | C-Index<sup>d</sup> |
| **Model 1** | 1.57<br>(1.38 to 1.77)<sup>c</sup> | NA | 0.688 | 1.51<br>(1.33 to 1.71) | NA | 0.674 |
| **Plus smoking** | 1.34<br>(1.19 to 1.52) | 35 | 0.704 | 1.36<br>(1.20 to 1.54) | 32 | 0.700 |
| **Plus**<br>**exercise<sup>e</sup>** | 1.56<br>(1.38 to 1.77) | 1 | 0.688 | 1.50<br>(1.33 to 1.70) | 10 | 0.668 |
| **Plus smoking and**<br>**exercise**<br>**linear model<sup>f</sup>** | 1.34<br>(1.18 to 1.52) | 35 | 0.701 | 1.36<br>(1.20 to 1.54) | 32 | 0.691 |
| **Plus**<br>**smoking and**<br>**exercise non-linear**<br>**model<sup>g</sup>** | 1.31<br>(1.15 to 1.49) | 40 | 0.706 | 1.28<br>(1.13 to 1.45) | 45 | 0.715 |

Abbreviations: HR, Hazard ratio; CI, confidence interval; NA data not applicable
<sup>a</sup>Of a total of 9775 participants there were 2480 deaths in the Whitehall study
<sup>b</sup>Percentage attenuation= $100 \times (\ln_{\text{Model 1+ health behaviours}} - \ln_{\text{Model 1}}) / (\ln_{\text{Model 1}})$ .
<sup>c</sup>Lowest socioeconomic position compared to highest position, adjusted for sex and age
<sup>d</sup>Concordance Index for model's performance on test data
<sup>e</sup>Participants were classified as either active (>2.5 hours/week of moderate exercise or >1.25 hours/week of vigorous exercise) or inactive (<2.5 hour/week of moderate exercise or <1.25 hour/week of vigorous exercise)
<sup>f</sup>Smoking and Exercise entered simultaneously into the cox model with age and sex (stcox i.grklump age sex smoke exercise)
<sup>g</sup>Smokers who don't exercise were classified as 3, smokers who exercise 2, non-smokers who don't exercise 1 and non-smokers who exercise 0.

## Discussion

The link between socioeconomic status and mortality can be traced back to 1929 when Edgar Sydenstricker released his report “Economic status and the incidence of illness” [20]. Most of the inequality in mortality associated with socioeconomic status can be statistically attributed to health behaviours. This could be due to the increased prevalence of detrimental health behaviours in lower socioeconomic groups [21].

Further, considering changes in exposure to certain health behaviours across follow-up was found to increase the explanatory power of the model. It has been suggested that those of a lower socioeconomic status are less willing/able to change health behaviours [22]. This would support our findings. The increase in explanatory power owing to longitudinal consideration seen for smoking was less than that seen for other health behaviours. This may be because smoking is a relatively invariant behaviour which exerts its adverse effects over a long period of adult life. Hence, time-varying data adds little new information for this.

The effect of reverse causation should also be considered-for instance, poorer health at earlier phases could lead to e.g.1. physical inactivity at later phases or e.g.2. inability to work and thus to afford what constitutes a healthy diet at later phases [23]. This partly explains why longitudinal consideration accounts for the gradient in mortality significantly more than baseline consideration. Further analysis could seek to understand these relationships for other morbidities that come with ageing, or other causes of death.

Adding more variables and building a more complex diet score increases further the explanatory power of the model. This was illustrated by a more detailed dietary analysis improving the ability of the model to account for the socioeconomic gradient. There is evidence that shows that dietary behaviours are poorer for those who are poorer [24], and this analysis shows that these behaviours are detrimental to lifespan.

At baseline there were fewer variables corresponding to diet compared with later phases, and we could not capture all these due to requiring the same variables at all phases. Nonetheless, further investigation making use of this could yield insightful results about the role of diet in mortality.

A nonlinear model was created between smoking and exercise, which found (through measuring the C-index) that by including the nonlinear relationship between exercise and smoking, the model’s ability to predict mortality improved. The C-index for the nonlinear model was 0.024 higher than the linear model showing that it performs better on a test set of data. Additionally, the nonlinear model had a 13% higher explanatory power when explaining the socioeconomic gradient compared to the linear model in data assessed longitudinally. This could be because smoking can limit someone’s ability to exercise. Participants from lower socioeconomic groups were found to be less active [25] and more likely to smoke [26]. The nonlinear model may suggest that smoking and exercise could come hand in hand.

### Limitations

There are no confidence intervals on the percentage attenuations achieved. This makes the results of the study more difficult to interpret because the confidence intervals provide information about whether differences observed are significant. One way that this could be resolved is to use bootstrapping.

Is Whitehall II representative of the wider population? It contains information pertaining to mostly male, mostly white, white-collar workers living in London and does not account representatively for those unemployed or living in poverty. However, it could be argued that with such a stark mortality gradient in one workforce, the corresponding gradient in the wider population would by deduction be as stark if not more so. Therefore, we hypothesise the validity of the study would not be affected.

Responding to questionnaires about health behaviours could lead to self-report bias like social desirability bias. Social pressure may lead to an unwillingness to admit to not having acted in a socially desirable manner. Self-administered questionnaires reduce susceptibility to information bias like social desirability bias; however, they are more prone to missing data on sensitive information.

Additionally, this study doesn’t enable us to draw conclusions on the relative importance of these selected health behaviours in comparison with other factors including psychosocial factors because this analysis wasn’t included. To improve this study, we could include psychosocial and financial factors into the models and compare their effects.

The proportional hazards assumption was taken. This implies that variables’ hazards remain constant throughout the course of the analysis. If this were not the case, then inferences of the analysis might be false: of detriment to the validity of the study. We are confident that the analysis does not violate the assumption, but to confirm this Schoenfeld residuals should be calculated.

Imputation with data from the previous phase for longitudinal analysis may lead to less representation of changes in health behaviours in this study. However, around 25% of the participants had data missing in the study, so removing these participants entirely as at baseline would mean loss of a large section of data. Therefore, it can be argued as being justified.

### Future implications

Our findings may have important public health implications. There is certainly a plausible causal link between the groups of factors studied (smoking, diet, exercise, alcohol consumption) and morbidity/mortality [27-30]. This implies that creating health policy interventions that focus on individual health behaviours could substantially reduce inequalities in health and improve the population’s health.

However, precise estimates of the importance of the risk factors studied will depend on the country and time period. Analytical methods can be improved to better understand the influences of health, across the population, and according to socioeconomic position. These methods can be applied to other types of risk factors such as autonomy at work and job security to further improve this study. Further implications for extending our work can include creating nonlinear relationships between other variables and incorporating further factors to obtain a richer picture of an individual’s health behaviours.

## Conclusion

This study suggests the importance of taking into account how health behaviours accumulate over time when considering their position in social inequalities. This study demonstrates that when adjusting for health behaviours (smoking, alcohol consumption, diet and physical activity) when assessed longitudinally with 32 years of follow-up, they can explain over 90% of the socioeconomic gradient in mortality compared to 51% for consideration solely at baseline. This proves that information on exposure at one point in the life course does not represent the whole life experiences of the population. A nonlinear model was also created between smoking and exercise. This explained 45% of the social inequality, showing the importance of incorporating and modelling potential non-linear interactions between variables. More complete diet indexes, which incorporate additional variables, also showed a very significant increase, with 91% of this inequality in mortality now being accounted for.

Different socioeconomic groups are subject to an array of social, cultural and economic influences over their life course, which shape their health behaviours, and how these change over time is also influenced by this array of factors. Together these findings indicate that if actionable factors are targeted by public health interventions and reversed, some of the social inequalities in mortality can be lessened. This may become increasingly critical as the cost-of-living crisis worsens.

## Data Availability

All data is protected in Dementias Platform UK and available upon application with reasonsable request.

https://portal.dementiasplatform.uk/

